# An exposome-wide assessment of 6600 SomaScan proteins with non-genetic factors in Chinese adults

**DOI:** 10.1101/2024.10.24.24316041

**Authors:** Ka Hung Chan, Jonathan Clarke, Maria G. Kakkoura, Andri Iona, Baihan Wang, Charlotte Clarke, Neil Wright, Pang Yao, Mohsen Mazidi, Pek Kei Im, Maryam Rahmati, Christiana Kartsonaki, Sam Morris, Hannah Fry, Iona Y Millwood, Robin G Walters, Yiping Chen, Huaidong Du, Ling Yang, Daniel Avery, Dan Valle Schmidt, Yongmei Liu, Canqing Yu, Dianjianyi Sun, Jun Lv, Michael Hill, Liming Li, Robert Clarke, Derrick A Bennett, Zhengming Chen, China Kadoorie Biobank Collaborative Group

## Abstract

**Background:** Proteomics offer new insights into human biology and disease aetiology. Previous studies have explored the associations of SomaScan proteins with multiple non-genetic factors, but they typically involved Europeans and a limited range of factors, with no evidence from East Asia populations.

**Methods:** We measured plasma levels of 6,597 unique human proteins using SomaScan platform in ∼2,000 participants in the China Kadoorie Biobank. Linear regression was used to examine the cross-sectional associations of 37 exposures across several different domains (e.g., socio-demographic, lifestyle, environmental, sample processing, reproductive factors, clinical measurements and frailty indices) with plasma concentrations of specific proteins, adjusting for potential confounders and multiple testing.

**Findings:** Overall 12 exposures were significantly associated with levels of >50 proteins, with sex (n=996), age (n=982), ambient temperature (n=802) and BMI (n=1035) showing the largest number of associations, followed by frailty indices (n=465) and clinical measurements (e.g., RPG, SBP), but not diet and physical activity which showed little associations. Many of these associations varied by sex, with a large number of age-related proteins in females also associated with menopausal status. Of the 6,597 proteins examined, 43% were associated with at least one exposure, with the proportion higher for high-abundance proteins, but certain biologically-important low-abundance proteins (e.g., PSA, HBD-4) were also associated with multiple exposures. The patterns of associations appeared generally similar to those with Olink proteins.

**Interpretation:** In Chinese adults an exposome-wide assessment of SomaScan proteins identified a large number of associations with exposures and health-related factors, informing future research and analytic strategies.

## Introduction

Proteins play essential roles in all living cells, and an optimal balance of protein levels influence human health. Previous studies of individual plasma proteins have identified many easily measured biomarkers relevant for disease diagnosis and understanding of disease mechanisms, including troponin reflecting cardiac injury,^1^ alanine transaminase (ALT) reflecting liver damage,^2^ and C-reactive protein (CRP) reflecting systemic inflammation.^3^ Recent advances in high-throughput proteomic assays now enable measurements of thousands of plasma proteins with high levels of accuracy.^4^ This permits a more comprehensive investigation on the molecular mechanisms underlying disease aetiology.^4^

An estimated 70-90% of disease risk in adults could be attributed to non-genetic risk factors, collectively known as the “exposome”,^5^ which act through multiple complex biological pathways in disease pathogenesis, with plasma proteins being likely intermediate markers of exposure or adverse effects. Recent population-based proteomics studies have assessed the associations of proteins, assayed using Olink or SomaScan platforms, with several well-established risk factors (e.g., adiposity, ageing, and smoking) and their associated biological processes.^6–10^ However, most previous studies were conducted in European populations,^7–10^ and did not provide systematic evaluations of impact of a wider spectrum of non-genetic factors on the plasma proteome.^11,12^ An “exposome-wide” study of the correlates of plasma protein levels in diverse populations can inform future research priorities and guide analytical approaches (e.g., adjustment for potential confounders when studying specific exposures).

We undertook comprehensive assessment of the associations of >7000 SomaScan proteins with 37 major non-genetic risk factors in ∼2000 Chinese adults in the China Kadoorie Biobank (CKB). The main objectives of the present study were to (1) systematically explore the exposure profiles of ∼7000 SomaScan proteins in 2000 Chinese adults; (2) assess the profiles in relation to normalised and non-normalised SomaScan protein levels; and (3) compare the findings with a parallel investigation^13^ involving ∼3000 proteins measured using an antibody-based Olink proteomics platform.

## Methods

### Study population and design

Details of the study design and participants characteristics of CKB have been reported elsewhere.^11^ Briefly, CKB recruited ∼512,000 adults aged 30-79 years from 10 geographically diverse areas in 2004-2008. At baseline, trained health workers administered a comprehensive laptop-based questionnaire and recorded physical measurements, including anthropometry and blood pressure, using regularly-calibrated instruments following standardised protocols. Desktop biochemical assays included random plasma glucose (RPG) and hepatitis B surface antigen (HBsAg). A 10-mL non-fasting (with time since the last meal recorded) blood sample was collected from each participant, and then processed and stored in liquid nitrogen. The present study involved 2,026 randomly selected subcohort participants who had no prior history of cardiovascular disease, originally sampled along with 1,951 cases of incident ischaemic heart disease (IHD) for a case-cohort study.^6,14^

The CKB was approved by the Ethical Review Committee of the Chinese Center for Disease Control and Prevention (Beijing, China) and the Oxford Tropical Research Ethics Committee, University of Oxford (Oxford, UK), and all participants provided written informed consent upon recruitment.

### Proteomic assays

Details of the proteomic assays in the CKB have been described elsewhere.^6,14,15^ In brief, 60µl plasma aliquots in 2D-barcoded microtubes of 3,977 participants were delivered to the Somalogic Laboratory in Colorado, USA for profiling using SomaScan Assay v4.1, which covers 7,596 slow off-rate modified aptamers (SOMAmers) as protein-binding reagents, with 7,289 SOMAmers targeting 6,597 human proteins. Samples were randomly aliquoted into 96-well plates (including 11 wells for external control samples, including 5 calibrator, 3 QC, and 3 buffer samples). The raw output of the SomaScan assay were standardised based on external control samples to account for variability in microarrays and variations within and between plates. With an optional procedure of adaptive normalisation by maximum likelihood (ANML) to an external reference, which controls for inter-sample variability, the results were provided in normalised and non-normalised values in relative fluorescence units (RFU).^16^ The output values were further natural log-transformed in the main analysis. The limit of detection (LOD) for SOMAmers were defined with external buffer samples. Quality control (QC) checks were conducted comparing the median of QC samples on each plate to the reference, with a cross-place QC indicator (pass/flag) assigned to each SOMAmer. The 7,289 SOMAmers targeting human proteins were mapped to proteins based on their UniProt IDs supplied by Somalogic. Details of individual proteins and their distributions by sex are shown in **eTable 1**.

### Selected baseline characteristics

From the baseline questionnaire and physical measurements, we identified 37 key characteristics from six broad categories: demographics (e.g., age, sex, study area), lifestyle (e.g., alcohol, smoking, diet), environmental factors (e.g., air pollution, ambient temperature), health and wellbeing (e.g., medical history and mental health), clinical measurements (e.g., BMI, HBV, RPG) and female reproductive factors (e.g., age at menarche, age at menopause, parity) (**eTable 2**). We also derived composite indices for (i) healthy lifestyle (ranging from 0 to 5, with higher score indicating a healthier lifestyle), based on our previous publications, which takes into account smoking, alcohol intake, physical activity, dietary habits, and body shape;^17–19^ and (ii) frailty, based on 28 variables on self-reported medical conditions, symptoms, signs, and physical measurements that capture different aspects of cumulative health status deficits as described previously^20^ (see derivation details in **eTable 2**).

### Statistical analysis

The present study used a comparable analytical strategy to that used for Olink proteomics in a parallel paper.^13^ From the original subcohort of 2,026 participants, we excluded individuals with missing SomaScan proteomics data (n=4), whose blood sample with hybridization control scale factor out of range (n=7), and those with missing ambient temperature data (n=20), leaving 1,998 sub-cohort participants (1243 females, 755 males) for the main overall and sex-specific analyses.

Selected baseline characteristics were examined by sex, directly standardised according to age and study areas of the original CKB population structure to facilitate comparison. Multivariable linear regression was used to examine the associations of the 37 baseline characteristics with normalized plasma protein levels (in RFU), adjusting for age, age^2^, sex, study area, fasting time, fasting time^2^, outdoor temperature, outdoor temperature^2^ and plate ID. For proteins targeted by multiple SOMAmers, the association showing the smallest p-values per exposure was selected. The analyses were repeated on non-normalized protein levels to evaluate the impact of ANML, which might have attenuated meaningful inter-individual heterogeneity. We also examined the associations (with normalised protein levels) by three classes of protein abundance, defined according to the SomaScan-supplied dilution factor for each protein, as low-(2 x 10^-1^), moderate- (5 x 10^-3^), and high-(5 x 10^-5^) abundance.

Given the high correlation between many SOMAmers (or protein levels), we adopted the multiple testing correction approach described by Gadd et al.^21^ A Bonferroni-adjusted p-value threshold was applied, based on 1,186 principal components explaining 90% of the cumulative variance in 7,335 SOMAmers (**eFigure 1** and **eTable 3**), and 21 components explaining 90% of the cumulative variance in the 32 exposures tested, plus 5 additional exposures for reproductive factors.^21^ This adjustment was applied across all linear regression models, with a Bonferroni-adjusted p-value threshold at 0.05/(1186 × 26) = 1.62 × 10^−6^. All statistical analyses were performed using R version 4.1.2^22^ and packages ‘*tidyverse’* and ‘*ggplot2’*.

## Results

**Table 1** shows the distribution of the 37 baseline characteristics among the 1998 participants included, overall and by sex. The mean age at baseline was 50.8 (SD 10.5) years and 62.2% were females. Males had higher prevalence of alcohol drinking (37.2% vs 2.6%) and smoking (63.3% vs 2.3%) than females, but similar prevalence of self-reported poor health and prior diseases, and slightly lower healthy lifestyle index.

**Table 1.**
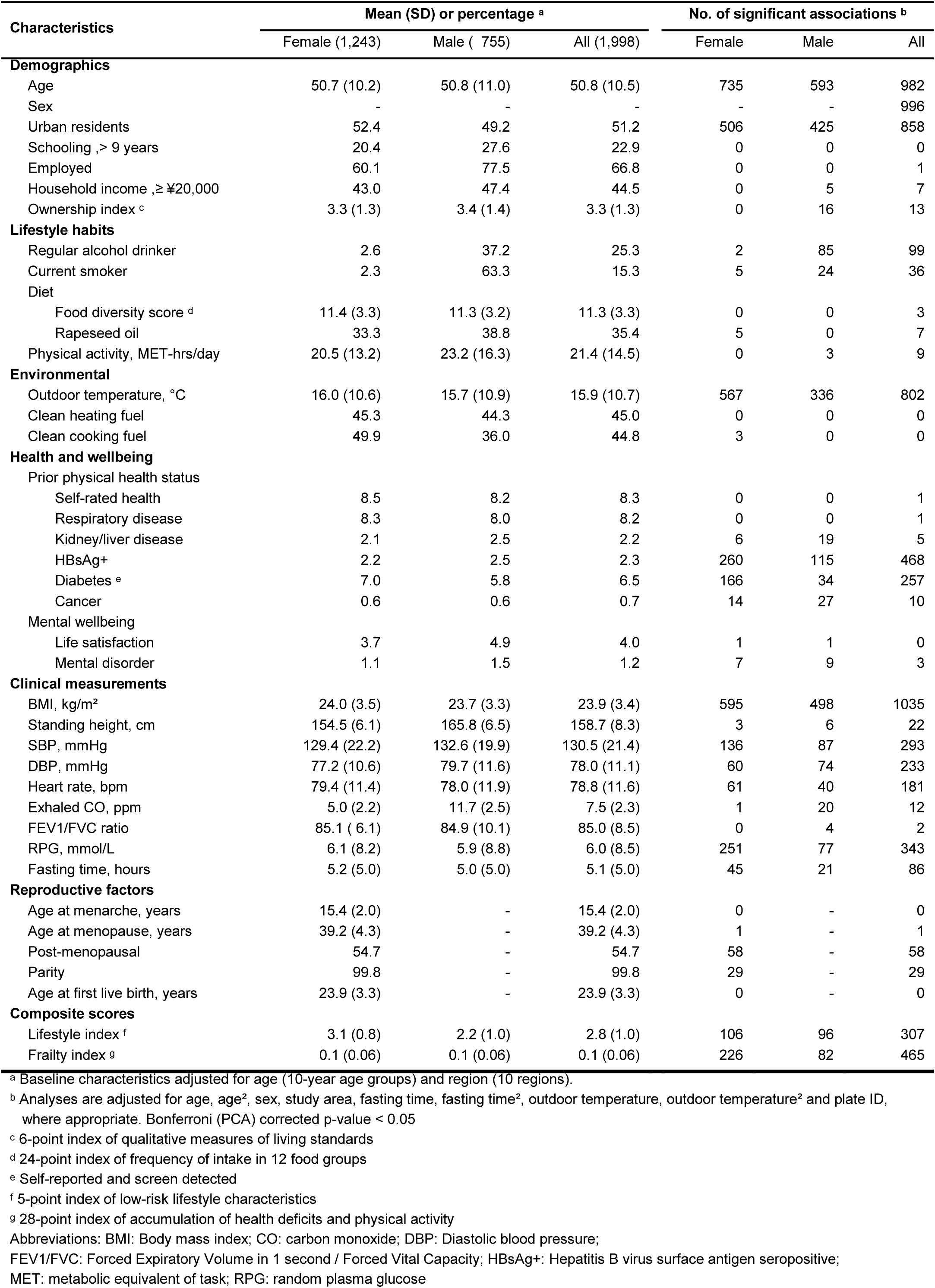
Baseline characteristics of participants and their associations with SomaScan protein biomarkers.

| Characteristics | Mean (SD) or percentage <sup>a</sup> |  |  | No. of significant associations <sup>b</sup> |  |  |
| --- | --- | --- | --- | --- | --- | --- |
|  | Female (1,243) | Male ( 755) | All (1,998) | Female | Male | All |
| **Demographics** |  |  |  |  |  |  |
| Age | 50.7 (10.2) | 50.8 (11.0) | 50.8 (10.5) | 735 | 593 | 982 |
| Sex | - | - | - | - | - | 996 |
| Urban residents | 52.4 | 49.2 | 51.2 | 506 | 425 | 858 |
| Schooling ,> 9 years | 20.4 | 27.6 | 22.9 | 0 | 0 | 0 |
| Employed | 60.1 | 77.5 | 66.8 | 0 | 0 | 1 |
| Household income ,≥ ¥20,000 | 43.0 | 47.4 | 44.5 | 0 | 5 | 7 |
| Ownership index <sup>c</sup> | 3.3 (1.3) | 3.4 (1.4) | 3.3 (1.3) | 0 | 16 | 13 |
| **Lifestyle habits** |  |  |  |  |  |  |
| Regular alcohol drinker | 2.6 | 37.2 | 25.3 | 2 | 85 | 99 |
| Current smoker | 2.3 | 63.3 | 15.3 | 5 | 24 | 36 |
| Diet |  |  |  |  |  |  |
| Food diversity score <sup>d</sup> | 11.4 (3.3) | 11.3 (3.2) | 11.3 (3.3) | 0 | 0 | 3 |
| Rapeseed oil | 33.3 | 38.8 | 35.4 | 5 | 0 | 7 |
| Physical activity, MET-hrs/day | 20.5 (13.2) | 23.2 (16.3) | 21.4 (14.5) | 0 | 3 | 9 |
| **Environmental** |  |  |  |  |  |  |
| Outdoor temperature, °C | 16.0 (10.6) | 15.7 (10.9) | 15.9 (10.7) | 567 | 336 | 802 |
| Clean heating fuel | 45.3 | 44.3 | 45.0 | 0 | 0 | 0 |
| Clean cooking fuel | 49.9 | 36.0 | 44.8 | 3 | 0 | 0 |
| **Health and wellbeing** |  |  |  |  |  |  |
| Prior physical health status |  |  |  |  |  |  |
| Self-rated health | 8.5 | 8.2 | 8.3 | 0 | 0 | 1 |
| Respiratory disease | 8.3 | 8.0 | 8.2 | 0 | 0 | 1 |
| Kidney/liver disease | 2.1 | 2.5 | 2.2 | 6 | 19 | 5 |
| HBsAg+ | 2.2 | 2.5 | 2.3 | 260 | 115 | 468 |
| Diabetes <sup>e</sup> | 7.0 | 5.8 | 6.5 | 166 | 34 | 257 |
| Cancer | 0.6 | 0.6 | 0.7 | 14 | 27 | 10 |
| Mental wellbeing |  |  |  |  |  |  |
| Life satisfaction | 3.7 | 4.9 | 4.0 | 1 | 1 | 0 |
| Mental disorder | 1.1 | 1.5 | 1.2 | 7 | 9 | 3 |
| **Clinical measurements** |  |  |  |  |  |  |
| BMI, kg/m<sup>2</sup> | 24.0 (3.5) | 23.7 (3.3) | 23.9 (3.4) | 595 | 498 | 1035 |
| Standing height, cm | 154.5 (6.1) | 165.8 (6.5) | 158.7 (8.3) | 3 | 6 | 22 |
| SBP, mmHg | 129.4 (22.2) | 132.6 (19.9) | 130.5 (21.4) | 136 | 87 | 293 |
| DBP, mmHg | 77.2 (10.6) | 79.7 (11.6) | 78.0 (11.1) | 60 | 74 | 233 |
| Heart rate, bpm | 79.4 (11.4) | 78.0 (11.9) | 78.8 (11.6) | 61 | 40 | 181 |
| Exhaled CO, ppm | 5.0 (2.2) | 11.7 (2.5) | 7.5 (2.3) | 1 | 20 | 12 |
| FEV1/FVC ratio | 85.1 ( 6.1) | 84.9 (10.1) | 85.0 (8.5) | 0 | 4 | 2 |
| RPG, mmol/L | 6.1 (8.2) | 5.9 (8.8) | 6.0 (8.5) | 251 | 77 | 343 |
| Fasting time, hours | 5.2 (5.0) | 5.0 (5.0) | 5.1 (5.0) | 45 | 21 | 86 |
| **Reproductive factors** |  |  |  |  |  |  |
| Age at menarche, years | 15.4 (2.0) | - | 15.4 (2.0) | 0 | - | 0 |
| Age at menopause, years | 39.2 (4.3) | - | 39.2 (4.3) | 1 | - | 1 |
| Post-menopausal | 54.7 | - | 54.7 | 58 | - | 58 |
| Parity | 99.8 | - | 99.8 | 29 | - | 29 |
| Age at first live birth, years | 23.9 (3.3) | - | 23.9 (3.3) | 0 | - | 0 |
| **Composite scores** |  |  |  |  |  |  |
| Lifestyle index <sup>f</sup> | 3.1 (0.8) | 2.2 (1.0) | 2.8 (1.0) | 106 | 96 | 307 |
| Frailty index <sup>g</sup> | 0.1 (0.06) | 0.1 (0.06) | 0.1 (0.06) | 226 | 82 | 465 |

<sup>a</sup> Baseline characteristics adjusted for age (10-year age groups) and region (10 regions).
<sup>b</sup> Analyses are adjusted for age, age<sup>2</sup>, sex, study area, fasting time, fasting time<sup>2</sup>, outdoor temperature, outdoor temperature<sup>2</sup> and plate ID, where appropriate. Bonferroni (PCA) corrected p-value < 0.05
<sup>c</sup> 6-point index of qualitative measures of living standards
<sup>d</sup> 24-point index of frequency of intake in 12 food groups
<sup>e</sup> Self-reported and screen detected
<sup>f</sup> 5-point index of low-risk lifestyle characteristics
<sup>g</sup> 28-point index of accumulation of health deficits and physical activity
Abbreviations: BMI: Body mass index; CO: carbon monoxide; DBP: Diastolic blood pressure;
FEV1/FVC: Forced Expiratory Volume in 1 second / Forced Vital Capacity; HBsAg+: Hepatitis B virus surface antigen seropositive;
MET: metabolic equivalent of task; RPG: random plasma glucose

Overall, 29 exposures were significantly associated with at least one protein after Bonferroni-adjustment, with 12 showing significant associations with >50 proteins (**Table 1**; **Figure 1**). In particular, sex (n=996), age (n=982), outdoor temperature (n=802), and BMI (n=1035) had the largest numbers of significant associations, followed by several clinical measurements (e.g. SBP and RPG), health and wellbeing indicators (e.g., HBsAg status and diabetes) and the lifestyle and frailty indices (**Table 1**; **Figure 1**). In sex-specific analyses, there were more significant associations in females than in males, except for smoking (n_female_=5; n_male_=24) and alcohol consumption (n_female_=2; n_male_=85), with three of the five female reproductive factors showing significant associations with 1∼58 proteins (**Table 1**). Across the 6,597 unique human proteins from the 7,289 SOMAmers examined, 43% were associated with at least one exposure, with IGFBP-2 (n=14), BGN (n=13), FIX (n=13), FIXab (n=13), and HSP 70 (n=13) associated with the greatest number of exposures, mostly with clinical measurements and demographic factors, overall and in sex-specific analyses (**Figure 2; eFigure 2**).

**Figure 1.**
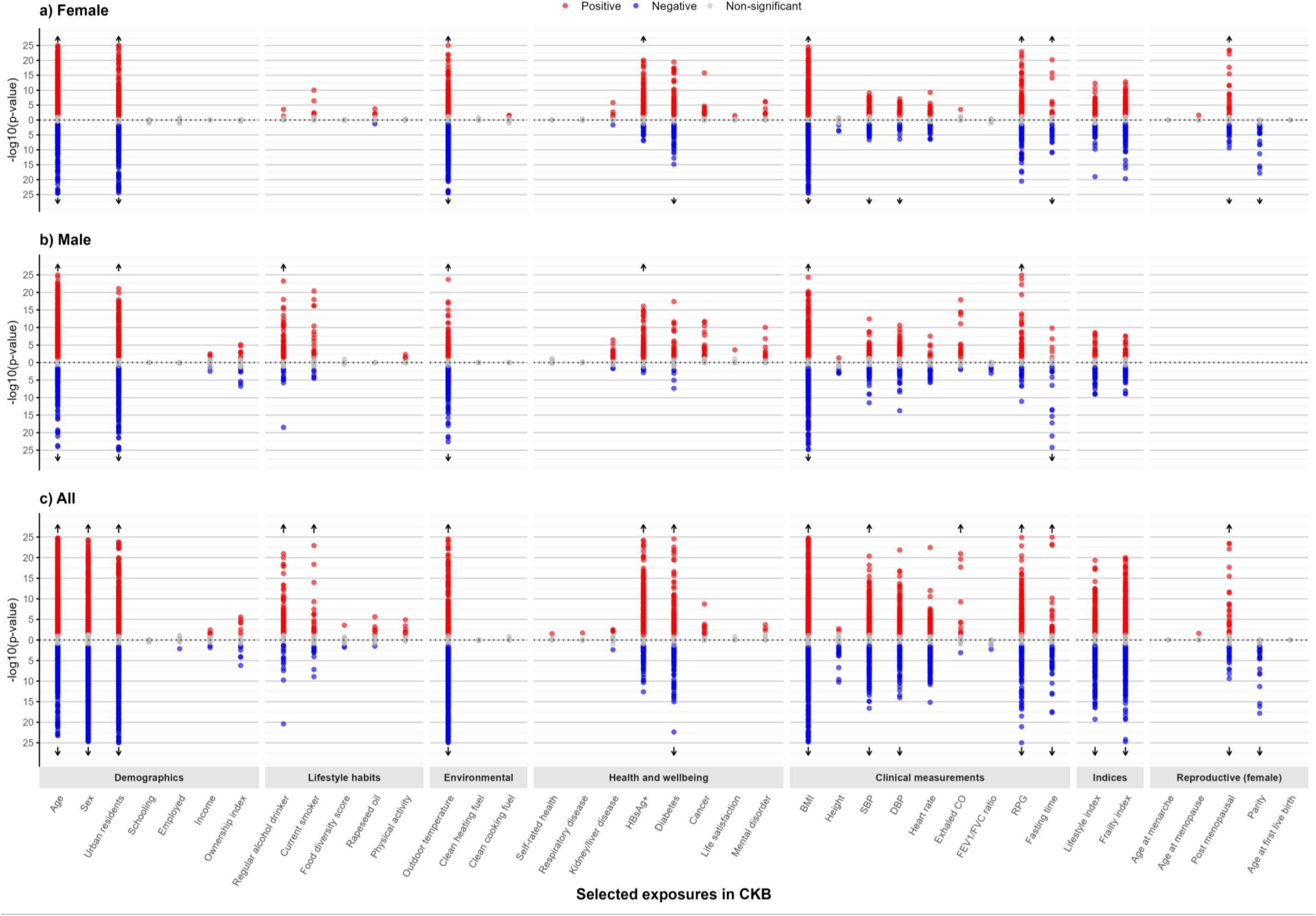
Exposure profile of 6597 SomaScan protein biomarkers in CKB, overall and by sex. Three Miami plots are presented: one for female-specific analysis, one for male-specific analysis, and one for overall analysis. The x-axis represents baseline characteristics grouped by category, while the y-axis shows the negative logarithm of the p-value (-log10 p-value) for the association between each exposure and protein biomarkers. Each dot represents the -log10 Bonferroni corrected p-value for these associations. For visualization purposes, -log10 p-values exceeding 25 are not displayed (indicated with arrow). Positive associations are shown in red, negative associations in blue, and non-significant associations in grey. Analyses are adjusted for age, age^2^, sex, study area, fasting time, fasting time^2^, outdoor temperature, outdoor temperature^2^ and plate ID, where appropriate. Abbreviations: BMI: Body mass index; CKB: China Kadoorie Biobank; CO: carbon-monoxide; DBP: Diastolic blood pressure; FEV1/FVC: Forced Expiratory Volume in 1 second / Forced Vital Capacity; HBV: Hepatitis B virus; MET: metabolic equivalent task; RPG: random plasma glucose

**Figure 2.**
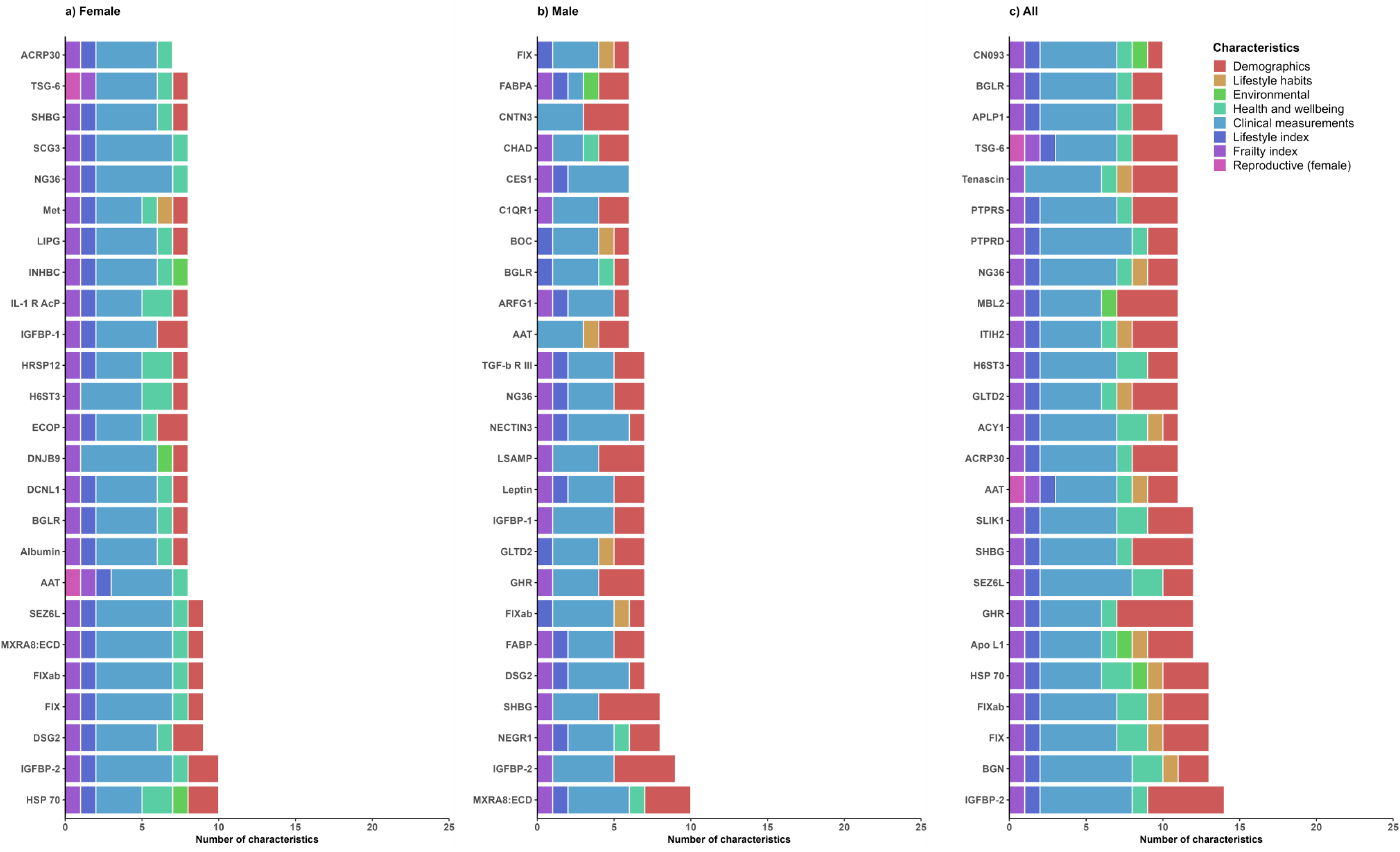
Exposure profiles of the top 25 SomaScan protein biomarkers with most associations, overall and by sex. The bar plots show the number of baseline associated with the 25 most frequently associated protein biomarkers after Bonferroni corrected p-value. The analyses are presented separately for females, males, and the overall. The x-axis represents the protein biomarkers, while the y-axis indicates the number of baseline characteristics associated with each protein. Bars are color-coded to represent different baseline characteristic groups. Analyses are adjusted for age, age^2^, sex, study area, fasting time, fasting time^2^, outdoor temperature, outdoor temperature^2^ and plate ID, where appropriate.

Of the 996 proteins associated with sex, the most significant associations included higher levels of leptin, FSH, and PZP in females and higher levels of PSA, HBD-4, and BPSA in males (**Figure 3ia**). Among the top 50 sex-related proteins, most were also associated with other exposures, particularly age (e.g., FSH, HCG) and BMI (e.g., leptin, FABP) (**Figure 3ib**). Importantly, there were 180 proteins uniquely associated with sex, but not with any other exposures (**eTable 4**). Furthermore, apart from alcohol drinking and smoking where the exposure-protein associations were stronger in females, most other associations appeared either comparable or slightly stronger in males (**eFigure 3**).

**Figure 3.**
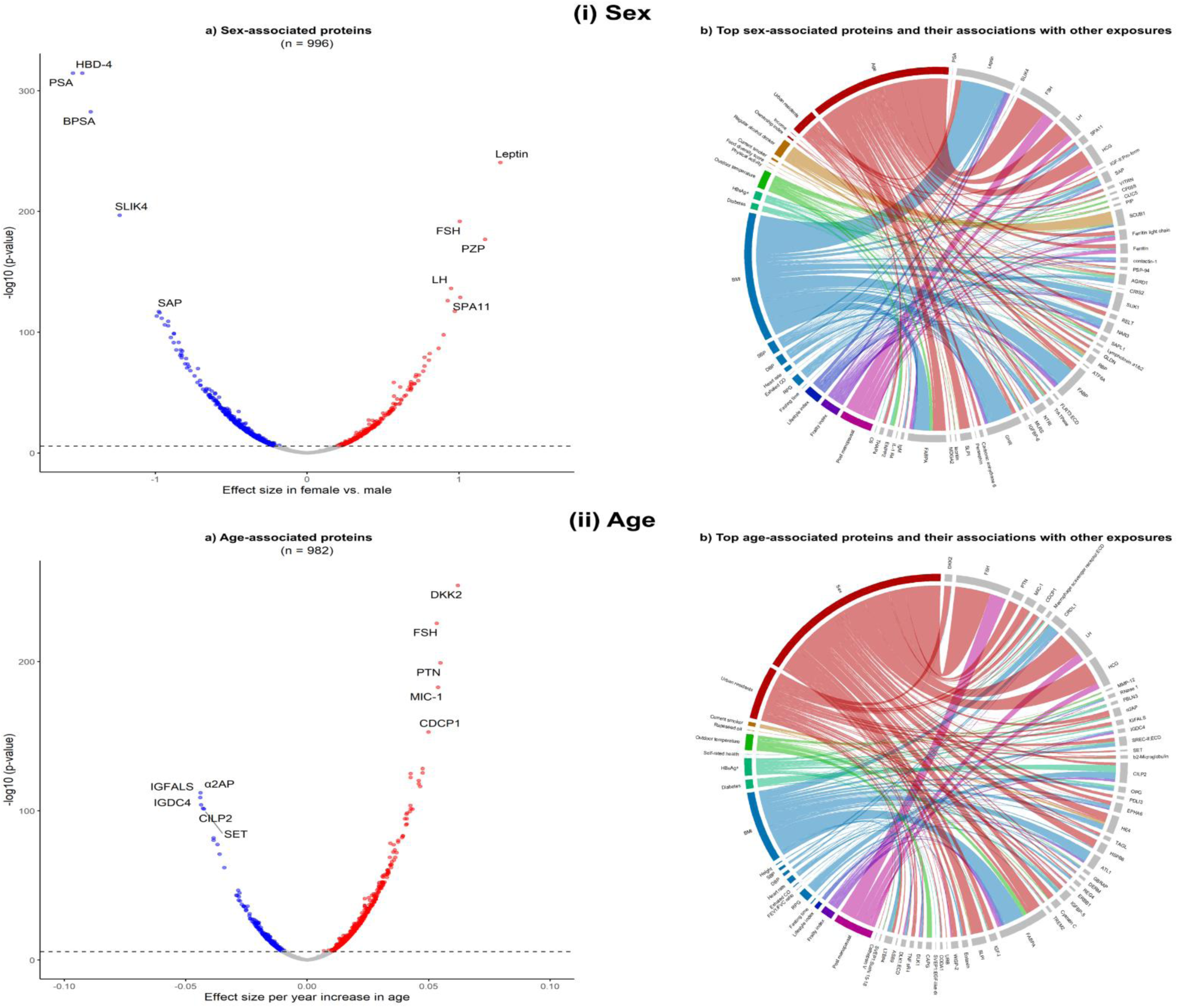
Sex- and age-associated SomaScan protein biomarkers and their associations with other exposures. Figures (i)a and (ii)a represent the associations of sex and age, respectively, with protein biomarkers. The x-axis represents the effect size of the association between sex or age and the protein biomarkers, while the y-axis indicates the –log10 p-value. Red dots denote positive Bonferroni corrected associations, blue dots denote negative Bonferroni corrected associations, and grey dots denote non-significant associations. Figures (i)b and (ii)b illustrate the top sex- and age-associated protein biomarkers, respectively, and their associations with other exposures. The width of the ribbons is inversely proportional to the p-value, indicating the strength of the association (smaller p-values correspond to wider ribbons). The colors of the ribbons represent different baseline characteristic groups. The top protein biomarkers that are not associated with other exposures are not presented in the figure. Analyses are adjusted for age, age^2^, sex, study area, fasting time, fasting time^2^, outdoor temperature, outdoor temperature^2^ and plate ID, where appropriate. Abbreviations: BMI: Body mass index; CO: carbon-monoxide; DBP: Diastolic blood pressure; HBV: Hepatitis B virus; RPG: random plasma glucose

As for the 982 proteins associated with age, DKK2, FSH, PTN, MIC-1, and CDCP1 showed the most statistically significant positive associations, whereas α2AP, IGFALS, IGDC4, CILP2, and SET showed the most significant inverse associations (**Figure 3iia**). Of the top 50 (by statistical significance) age-related proteins, most were also associated with other exposures, particularly sex (e.g., FSH, PTN), BMI (e.g., CRDL1, FABPA), and urban residence (e.g., DKK2, MIC-1) (**Figure 3iib**). In sex-specific analyses, age was associated with 735 and 593 proteins in females and males, respectively (**Table 1**), with most of the 357 overlapping proteins showing concordant associations (**eFigure 4**). Among the top age-associated proteins by sex, many were also associated with post-menopausal status, BMI, and urban residence in females, and with BMI and urban residence in males (**eFigure 5**). Consistently, the majority of the 58 proteins associated with post-menopausal status were also associated with age (e.g., FSH, LH, CRDL2, HCG), with some also being associated with BMI (e.g., FABPA, FABP) (**eFigure 6**).

HBsAg status and prevalent diabetes were associated with 468 and 257 proteins, respectively, but other health and wellbeing indicators, including self-rated health or other prior medical history showed few significant associations (**Table 1**). For HBsAg status, the top positively associated proteins were DHI1, NUD16, DJB12, C1QTNF3, and AKR1D1, while the top inversely associated proteins were CFHR5, SAP, RBP, IL-1 R AcP, and α2AP (**eFigure 7**). For prevalent diabetes, the top positively associated proteins included PLXB2 and SEMA6A and top inversely associated proteins were CILP2 and MXRA8:ECD (**eFigure 7**).

Several clinical measurements, including BMI, SBP, DBP, heart rate, and RPG (but not height, exhaled CO or lung function) were strongly associated with multiple proteins (**Table 1**). BMI had the highest number of associations (n=1035) among all exposures examined, with leptin, GHR, FABP, FABPA, and GPDA being the top positively associated proteins and IGFBP-2, IGFBP-1, WFKN2, SHBG, and SEZ6L being the top inversely associated proteins (**eFigure 8**). In the sex-specific analyses, leptin, GHR, FABP, FABPA, IGFBP-2, WFKN2, and SHBG were also top hits with BMI, with the same direction of association in males and females (**eFigure 9**). The proteins that had the most significant positive associations with SBP were GHR, INHBC, FIXab, FIX, and leptin, while those with the most significant inverse associations were renin, IGFBP-2, SHBG, H6ST2, and SCG3 (**eFigure 7**). Among the top RPG-associated proteins, there were positive associations with PLXB2, SEMA6A, SEM4D, SEM6B, and NFASC and inverse associatons with CILP2, MXRA8:ECD, COL15A1, SCG3, and ALB (**eFigure 7**).

Among the 307 proteins associated with healthy lifestyle index (**Table 1**), the most significant positive associations included TINAL, NCAM1, and NCAM-120, and the inverse associations included leptin, GPDA, and UGDH (**Figure 4ia**). The index-protein associations, as illustrated in the most strongly associated proteins, appeared to overlap with those with BMI and sex, followed by clinical measurements (e.g., SBP, DBP, RPG), and urban residence (**Figure 4ib**). The composite frailty index was associated with 465 proteins overall, and 226 and 82 in females and males, respectively (**Table 1**; **Figure 4iia**). Among the leading frailty-associated proteins, there were positive associations with CRP, ESPN, and HTRA1 and inverse associations with MXRA8:ECD, ANTR2, and SHBG (**Figure 4iia**). Importantly, the proteins most strongly associated with frailty index overlapped with most clinical measurements (particularly BMI), age, sex, and lifestyle index, in a largely coherent manner (i.e., opposing direction of association) as expected (**Figure 4iib**).

**Figure 4.**
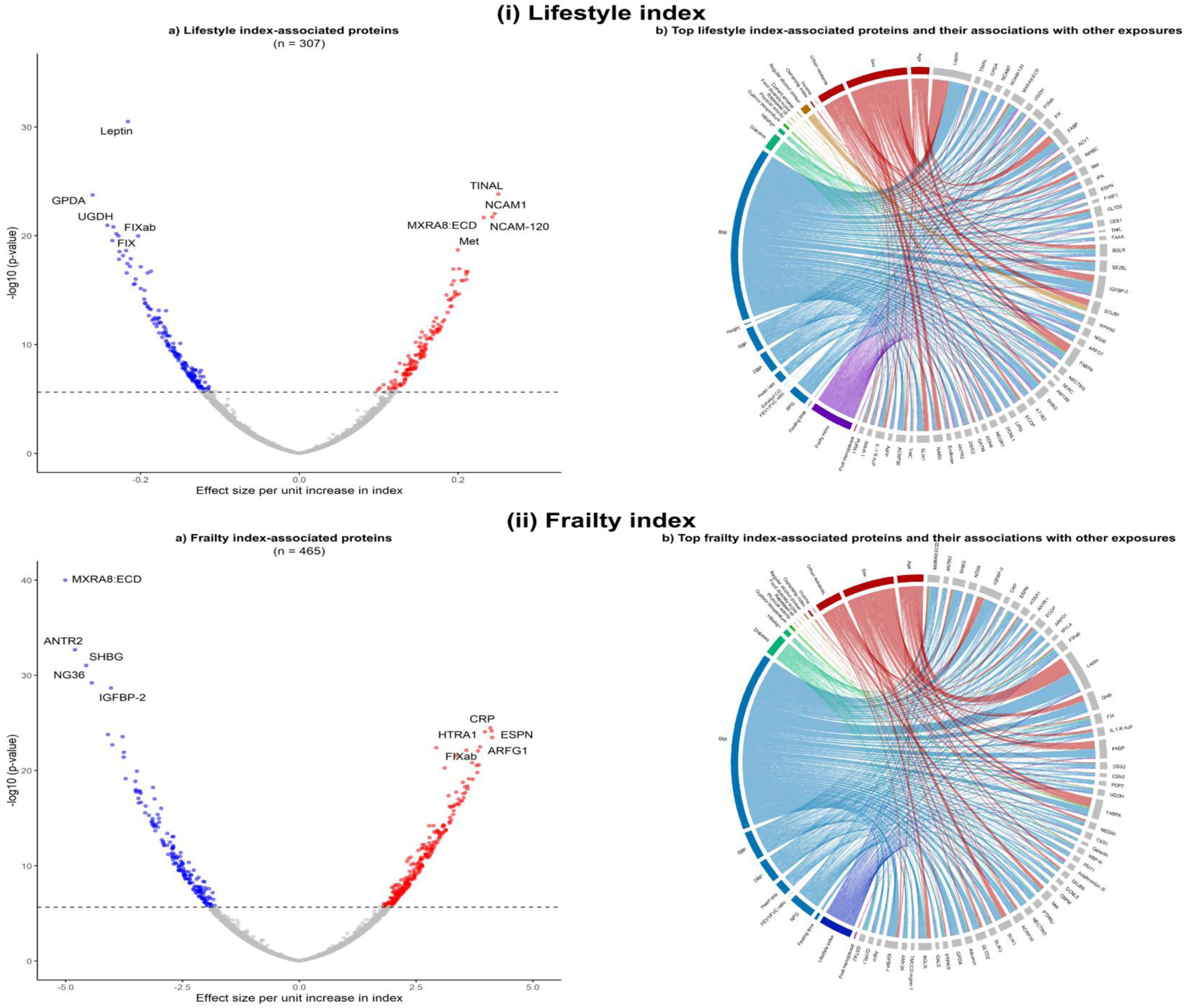
Lifestyle and frailty indices-associated SomaScan protein biomarkers and their associations with other exposures. Same as Figure 3.

Notably, we observed a general trend of higher proportion of significant associations with proteins of higher abundance (e.g., 41% proteins of high-abundance vs. 10% of low-abundance were associated with age), except for outdoor temperature (∼10% for both high- and low-abundance proteins) (**eTable 5; eFigure 10**). Analysis of non-normalised protein levels showed greater number (by 1.5 to 3 times) of significant associations with age, diabetes, BMI, SBP, DBP, heart rate, RPG, menopausal status, and lifestyle and frailty indices, but considerably fewer (15% lower) significant hits with outdoor temperature (**eTable 6; eFigure 11**). Generally, the proportional increment of significant associations in the non-normalised proteins were more prominent in females than males, except for BMI and frailty index (**eTable 6**). Importantly, the effect sizes of associations related to non-normalised protein levels were smaller than for normalised levels for age, BMI, SBP, RPG, prevalent diabetes, and frailty index, but the converse was true for lifestyle index, whereas the associations with sex were highly consistent (**eFigure 12**). Across the two parallel analyses, both SomaScan and Olink platforms yielded comparable numbers of significant proteomic associations with the multiple exposures, particularly with sex, age, alcohol drinking, smoking, HBsAg status, diabetes, clinical traits, menopausal status, healthy lifestyle and frailty indices (**eFigure 13**).

## Discussion

This exposome-wide analyses of almost 6,600 SomaScan proteins in ∼2,000 Chinese adults demonstrated significant associations of several major non-genetic risk factors with plasma levels of specific proteins. Exposures such as age, sex, ambient temperature, and BMI demonstrated the largest number of significant associations with multiple proteins, with many of these associations varying by sex. Other exposures including demographic factors, lifestyle habits, clinical measurements, and lifestyle and frailty indices were also related to a very large number of proteins. Overall, there was a larger proportion of significant associations with high-abundance proteins, but we still found associations with low-abundance but biological-important proteins not captured in the Olink immunoassays (e.g., PSA, HBD-4, DKK2).

The two parallel analyses across the SomaScan and Olink^13^ platforms yielded largely consistent findings, especially on factors associated with large number of proteins. Notably, the patterns of associations with Olink proteins were more similar to those for the non-normalised SomaScan proteins, consistent with previous investigations.^15^ Moreover, both platforms showed directionally consistent associations, as demonstrated for instance by leptin (higher in females in both platforms) and IGFBP-2 (inversely associated with frailty index in both platforms). Furthermore, a number of proteins that are included in the SomaScan platform but not in the Olink platform were significantly associated with many exposures (e.g. age positively associated with DKK2, and frailty index inversely associated with ANTR2), demonstrating the complementary advantages of the two proteomic platforms.

Previous studies have investigated the SomaScan plasma proteomic profiles of various exposures.^23–28^ However, they included primarily Western populations, used the earlier versions of the SomaScan platforms with fewer proteins measured, and typically focused on single or a small group of individual exposures.^23–27^ The only recent broad-spectrum study that used an earlier SomaScan (v4.0) platform to explore the genetic and non-genetic predictors of 4,775 plasma proteins measured in ∼8000 European adults, showed that non-modifiable factors, including genetic factors, age, and sex were the major determinants of plasma proteins (of 3,242 protein targets), which is somewhat in line with our findings (on age and sex).^28^ However, age in this European study was associated with a higher proportion of proteins than in our study,^28^ potentially because of the larger sample size. A few studies also used the earlier Olink platform assays to investigate the exposure profiles of ∼90 proteins.^29–31^ Our study is one of the first and largest proteomic-exposome profiles studies in China and East Asia that systematically evaluates the impact of a much wider spectrum of exposures on the plasma proteome. Our analyses also identified important differences in proteomic-exposome profiles between males and females, including many proteins known to be involved in sex-specific biological processes. For example, FSH that stimulates the growth and development of follicles in females,^32^ was higher in females than males, consistent with findings in the parallel CKB Olink proteomics study.^13^ Furthermore, levels of PZP, an immunosuppressive protein expressed by the placenta,^33^ were higher in females than in males, while the levels of PSA/BPSA and HBD-4 , which are expressed in prostate ^34^ and testes,^35^ respectively, were higher in males. We also found leptin, which is known to play a key role in regulating energy balance and controlling body weight,^36^ to be significantly higher in females, reflecting the sex-specific variations in body composition measurements,^37^ as observed in the parallel CKB Olink proteomics study.^13^ Interestingly, adjustment for sex in our analyses did not attenuate the associations of the sex-related proteins with other exposures, possibly reflecting other sex-independent effects of the proteins. For example, FSH was also associated with age and leptin was associated with BMI, independent of sex. Therefore, these proteins might also be involved in biological processes (metabolic, inflammatory, or ageing processes) that could be influenced by exposures like age and BMI,^38^ beyond their associations with sex. Genetic regulation of the plasma proteome has been previously reported to be largely similar among the two sexes,^39^ even though levels of plasma proteins were largely different between men and women, suggesting that these differences might be related mainly to differential non-genetic factor between the sexes,^40^ and highlighting the importance of performing sex-specific and sex-adjusted analyses in observational proteomic studies.

Importantly, some proteomic associations with individual lifestyle factors differed by sex. For example, TBG and sICAM-5, which have been previously associated with alcohol intake and smoking,^23,41^ respectively, were more strongly related to the corresponding exposures in males than in females in our study. These observed sex-differences likely reflect the markedly higher prevalence (and intensity) of alcohol drinking and smoking in men than in women in the Chinese population,^42,43^ which is also consistent with the higher number of significant hits with alcohol drinking and smoking in males. Interestingly, there were limited proteomic associations with the other individual lifestyle factors, most notably diet and physical activity, which is consistent with the previous SomaScan proteomics-exposome study in Europeans.^28^ Lifestyle factors could influence the proteome dynamically and these dynamic protein changes are challenging to be captured fully by cross-sectional snapshots of protein levels.^44^

Age was strongly and positively associated with DKK2, which is a glycoprotein known to regulate vertebrate development and to modulate the Wnt signalling pathway.^45^ DKK2 has been found to be involved in the development of several ageing-related diseases, including cancer.^46–48^ We also found older age to be strongly associated with higher levels of PTN, a secreted growth factor that regulates cellular proliferation, growth, differentiation and migration.^49^ Previous studies also reported higher levels of plasma PTN being significantly associated with chronological age.^24,50^ Additionally, the most strongly age-associated protein was α2AP, a serine protease inhibitor responsible for inactivating plasmin. Circulating plasmin is released in the plasma during fibrinolysis, which is an important regulator of blood coagulation process, and congenital deficiency of α2AP causes a rare bleeding disorder due to increased fibrinolysis,^51^ and an increased bleeding tendency with age in patients with heterozygous deficiency.^52^ We also found that many other age-related proteins were also independently associated with other risk factors. For example, CRDL1, a bone morphogenetic protein-4 antagonist, was associated with BMI, SBP and DBP. Moreover, in consistent with Olink proteins, we found numerous sex-specific top protein hits for age to be also associated with postmenopausal status, including FSH, which is known to be higher in menopausal women.^53^

We found a large number of proteins associated with several clinical as well as health and wellbeing-related traits. BMI was associated with the largest number of significant associations with proteins, which aligns with the relatively high protein variance attributable to measures of body composition reported previously in Europeans.^28^ A number of these associations were previously evident in both observational and genetic analyses, such as the positive association with leptin and FABP, which is involved in the uptake, metabolism and transport of long-chain fatty acids.^6,26^ Future studies should also consider assessing the associations of SomaScan proteins with central adiposity (e.g. waist circumference, waist-to-hip ratio), as it is known to be more strongly associated with risk of cardio-metabolic disease^54,55^ and with Olink proteins,^56^ compared with BMI. Prevalent diabetes and RPG were both positively associated with PLXB2 (receptor for semaphorins involved in cell-cell signalling) and inversely associated with CILP2 (involved in cartilage scaffolding), which are supported by prior observational ^57,58^ and genetic analyses.^27,59^ In addition to many new findings that need to be explored further in subsequent studies, we also identified some previously reported associations, such as SBP with renin (enzyme involved in the renin-angiotensin-aldosterone system)^60^ in both SomaScan and Olink studies, and HBV infection with SAP (part of the innate immune humoral arm)^61^.

The present study demonstrated a large number of proteomic associations with both the frailty ^20^ and healthy lifestyle indices.^17,18^ The two indices capture essentially opposing dimensions of health status, and many proteins showed opposing associations across the two indices, including HTRA1, MXRA8:ECD, ANTR2, leptin, GPDA, UGDH, IGFBP-2, and SHBG. HTRA1 is implicated in inhibiting active TGF-β, which is an anti-inflammatory cytokine,^62^ while MXRA8:ECD and ANTR2 are both involved in angiogenesis.^25^ GPDA was previously shown to be positively associated with current smoking and alcohol drinking in the Framingham Heart Study,^23^ which is line with our finding of an inverse association of lifestyle index with levels of this protein. Both GPDA and UGDH have also been implicated in multiple cancer types, and considered as possible therapeutic targets for treatment of cancer.^63,64^ In addition, consistent with our findings related to the two indices, circulating IGFBP-2 and SHBG were reported to be inversely associated with obesity,^65,66^ while a previous study showed lower circulating levels of IGFBP-3 (same protein family with IGFBP-2) to be associated with current smoking.^67^ Both IGFBP-2 and SHBG are involved in cancer and metabolic/reproductive system disorders, respectively, and they have also been identified in the Olink CKB study. Many indices-related proteins were also associated with individual exposures, particularly BMI, other clinical measurements, sex, and age, which is an expected finding as a number of these exposures are constituents of the indices.

Among the many exposures examined previously, the impact of outdoor temperature on the plasma proteome has been understudied.^29,68^ We found a large number novel significant associations that warrant further investigation. For example, SPINK6, which showed a strong positive association with ambient temperature, is a potent inhibitor of serine proteases that are essential for influenza A viruses infection in the airways.^69^ Similarly, LRP1 and MMP7, which are inversely associated with temperature, play important roles in lipid metabolism.^70,71^ However, we found fewer significant proteomic associations of proteins with several other exposures, including household air pollution, prevalent diseases (except diabetes), mental health, height, lung function and most reproductive factors. Future proteomic studies with a larger sample size and increased proteome coverage might be needed to further clarify the relevance of the observed associations and to identify new associations.^44^

For reasons that are not fully understood, we found that use of non-normalised proteomics panels yielded an even larger number of significant associations with several major exposures (e.g., age, diabetes, and frailty and healthy lifestyle indices). This is consistent with the previous studies conducted in CKB^15^ and with some previous studies using different approaches to proteomic profiling.^72–74^ The normalization procedure adopted by SomaLogic standardises the overall signal of each sample to an external reference to improve data quality.^75^ However, this process may reduce true extreme signals that are present in the general population, attenuating the number of the observed significant associations.^72,76^ Interestingly, while most associations with the two types of data were directionally consistent, the effect sizes of the overlapping associations were smaller for non-normalised than for normalised protein levels.

The present study is the first and largest study in East Asia that measured the levels of 6,600 plasma proteins using the SomaScan platform and conducted a comprehensive investigation of the proteomic associations with >35 exposures across different domains. This type of “exposome-wide” investigation, in parallel with that on 2923 Olink proteins,^13^ is also important to inform future analytical approaches (e.g. confounding adjustment) when examining specific exposures. Moreover, the SomaScan platform used in CKB captured a large number of low-abundance proteins not available in the Olink panels, which may be equally important for understanding biological pathways. Comprehensive analysis of both high- and low-abundance proteins contributes to a more complete map of biological processes, providing insights into disease pathogenesis and progression. However, the current study also had several limitations. First, there were more females than males, which might bias towards finding a larger number of significant associations in females. Moreover, there was limited statistical power for performing further subgroup analyses, besides the sex-specific analyses. Second, the present study was restricted to proteins measured only in plasma and to proteins that cover a modest proportion of the human proteome. Third, replication of our findings in external and independent cohorts was not possible, as currently there are limited available SomaScan proteomic data in other East Asian populations. Nevertheless, our findings are broadly consistent with the parallel analyses on Olink proteomics which included 2,168 proteins also targeted by SomaScan.^13^ Fourth, the direction of the associations studied herein could not be confirmed due to the cross-sectional nature of the study. Finally, although the analyses were adjusted for a range of key covariates, such as age, sex, region, and fasting time, residual confounding cannot be fully excluded. Further genetic investigations could allow us to identify potential causal associations between various exposures and proteins.

Overall, this large proteomic study in the Chinese population showed that several exposures, particularly, age, sex, ambient temperature, BMI and composite indices of healthy lifestyle and frailty were associated with a large number of proteins that are involved in multiple pathways. The associations between major exposures and proteins also varied by sex, potentially due to sex-specific biology and sex-differential lifestyles. The findings of the present study may inform priorities for future proteomics research and further studies are warranted to replicate the current findings in independent populations and to provide information on the causal relevance of these associations.

## Acknowledgements

The chief acknowledgment is to the participants, China Kadoorie Biobank project staff, staff of the China CDC and its regional offices for access to death and disease registries. The Chinese National Health Insurance scheme provided electronic linkage to all hospital admissions. The China Kadoorie Biobank study is jointly coordinated by the University of Oxford and the Chinese Academy of Medical Sciences.

## Open Access Statement

This research was funded in whole, or in part, by the Wellcome Trust [212946/Z/18/Z, 202922/Z/16/Z, 104085/Z/14/Z, 088158/Z/09/Z]. For the purpose of Open Access, the author has applied a CC-BY public copyright license to any Author Accepted Manuscript version arising from this submission.

## Ethics approval

The China Kadoorie Biobank (CKB) complies with all the required ethical standards for medical research on human subjects. Ethical approvals were granted and have been maintained by the relevant institutional ethical research committees in the UK and China.

## Consent to participate/publication

All participants provided written informed consent.

## Author Contributions

KHC, JC, MK, DAB, and ZC conceived and designed the study. JC conducted the statistical analyses and KHC and MK wrote the first draft of the manuscript.LL, and ZC as the members of CKB Steering Committee, designed and supervised the overall conduct of the study, including obtaining funding for the study. All other authors provided critical revision to the manuscript for important intellectual content. KHC, JC, MK, DAB and ZC are the guarantors of this work and take responsibility for the integrity and accuracy of the data analysis. DAB and ZC supervised the work. All authors contributed to the review and edit of the manuscript.

## Data Availability

Data from baseline, first and second resurveys, and disease follow-up are available under the CKB Open Access Data Policy to bona fide researchers. Sharing of genotyping data is constrained by the Administrative Regulations on Human Genetic Resources of the People’s Republic of China. Access to these and certain other data is available through collaboration with CKB researchers. Details of the CKB Data Sharing Policy are available at www.ckbiobank.org.

## Abbreviations

AKR1D1: 3-oxo-5-beta-steroid 4-dehydrogenase
ALT: Alanine aminotransferase
ANML: Adaptive normalisation by maximum likelihood
ANTR2: Anthrax toxin receptor 2
APOF: Apolipoprotein F
BGN: Biglycan
BMI: Body mass index
BPSA: Benign prostatic-specific antigen
C1QTNF1: C1q and tumor necrosis factor related protein 1
C1QTNF3: C1q and tumor necrosis factor related protein 3
CFHR5: Complement factor H-related 5
CILP2: Cartilage intermediate layer protein 2
CKB: China Kadoorie Biobank
CO: Carbon monoxide
CRDL1: Chordin-like protein 1
CRDL2: Chordin-like protein 2
CRP: C-reactive protein
DBP: Vitamin D-binding protein
DKK2: Dickkopf-related protein 2
DJB12: DnaJ homolog subfamily B member 12
ESPN: Espin
FABP: Fatty acid binding protein
FABPA: Fatty acid binding protein, adipocyte
FIX: Coagulation factor IX
FIXab: Coagulation factor IXab
FSH: Follicle stimulating hormone
GHR: Growth hormone receptor
GPDA: Glycerol-3-phosphate dehydrogenase [NAD(+)], cytoplasmic
HBD-4: Hemoglobin subunit delta-4
HBsAg: Hepatitis B surface antigen
HBV: Hepatitis B virus
HCG: Human chorionic gonadotropin
HTRA1: Htra Serine Peptidase 1
H6ST2: Heparan-sulfate 6-O-sulfotransferase 2
DHI1: Corticosteroid 11-beta-dehydrogenase isozyme 1
IGDC4: Immunoglobulin superfamily, dcc subgroup member 4
IGFALS: Insulin-like growth factor binding protein, acid labile subunit
IGFBP-2: Insulin-like growth factor binding protein 2
IGFBP-3: Insulin-like growth factor binding protein 3
IHD: Ischemic heart disease
IL-1: R AcP Interleukin-1 Receptor accessory protein
INHBC: Inhibin beta C chain
LH: Luteinizing hormone
LOD: Limit of detection
LPL: Lipoprotein lipase
LRP1: Ldl receptor related protein 1
MIC-1: Macrophage inhibitory cytokine 1
MMP7: Matrix metallopeptidase 7
MXRA8: Matrix remodeling associated 8
MXRA8:ECD: Matrix remodeling associated 8, extracellular domain
NCAM1: Neural cell adhesion molecule 1
NCAM-120: Neural Cell Adhesion Molecule 120 kda Isoform
NFASC: Neurofascin
NUD16: U8 snorna-decapping enzyme
PLXB2: Plexin B2
PSA: Prostate-specific antigen
PTN: Pleiotrophin
PZP: Pregnancy zone protein
QC: Quality control
RBP: Retinol-binding protein 4
RFU: Relative fluorescence units
RIDA: Rectal intestinal domain antigen
RPG: Random plasma glucose
SAP: Serum amyloid P-component
SBP: Systolic blood pressure
SCG3: Secretogranin-3
SEMA6A: Semaphorin-6A
SEM4D: Semaphorin-4D
SEM6B: Semaphorin-6B
SHBG: Sex hormone-binding globulin
sICAM-5: Soluble intercellular adhesion molecule 5
SOMAmer: Slow off-rate modified aptamers
SPINK6: Serine peptidase inhibitor, kazal type 6
TBG: Thyroxine-binding globulin
TINAL: Tubulointerstitial nephritis antigen-like 1
TTR: Transthyretin
UGDH: Udp-glucose 6-dehydrogenase
WFKN2: WAP, Kazal, immunoglobulin, Kunitz and NTR domain-containing protein 2
α2AP: A2-antiplasmin

